# Does coaching in the 3^rd^ year of medical school affect stress, anxiety and depression?

**DOI:** 10.1101/2021.07.13.21260303

**Authors:** Elizabeth Ross, Timothy Faw, Kyle Covington

## Abstract

**Background:** Medical students experience significant stress and anxiety during undergraduate education. Coaching is a possible way of supporting these students throughout this challenging time. To assess the benefits of coaching for medical students, a pilot study providing coaching was performed. This pilot assessed how coaching affected the mental health of medical students and how coaching was received by them.

**Methods:** Twelve third-year medical students were each given eight 30-60 minute coaching sessions. Each participant took the Hospital Anxiety and Depression Scale (HADS) and the Perceived Stress Scale (PSS) pre-, mid-and post-coaching. After coaching, there were three open-ended questions to measure the reactions to coaching and a scale to determine the likelihood of accessing coaching in the future.

**Results:** There was a significant effect of coaching on perceived stress(p=.023); a trend toward significant effect of coaching on anxiety(p=.057); and no effect of coaching on depression. Qualitative analysis indicated Affective responses (gaining perspective and self-awareness); Cognitive responses (goal setting and working through solving problems); and Skills responses (developing reflection abilities and critical thinking). Attributes of coaching included perceiving coaching as a positive, individualized and supportive experience that students were highly likely to access again.

**Conclusions:** Coaching holds promise as an intervention offered to medical students to reduce stress and anxiety, and provide positive support for students, preparing them for their professional futures.

**Key Points:**

1. Appreciate the stress and anxiety that medical students manage in medical school.
2. Coaching is an option that holds promise to support medical students.
3. Coaching has possible long-term outcomes that could help future physicians navigate their professional paths more effectively.
4. Students value individualized, positive support during medical school

## Background

Medical students in their undergraduate education may experience significant stress and anxiety because of the transitions and identity formation they are experiencing (Dyrbye *et al*.,2006)(Kumar *et al*.,2019). They are engaged in coursework and decisions about clerkships, sub-internships, applications for residency, and interviews, critical for their future professional and personal development. These students need to be supported in navigating this journey, make the best decisions for themselves and develop skills in finding their own most meaningful paths. Coaching is a way of supporting students throughout this tumultuous time while providing an opportunity for the development of agency, confidence and thoughtful decision-making as they assume their professional roles.

Coaching has been a mainstay in the executive environment, yet it has been slow to become integrated in academia. There have been efforts in recent years to offer coaching to faculty, staff and students entering their undergraduate years and coaching has emerged in graduate programs, such as medical school(Cary *et al*.,2019)(Dieorio and Miller,2016)(Dieorio and Hammoud, 2019). The delivery of coaching for medical students seems to exist in pockets and frequently the model and format are not clearly articulated, yet there has been a trend toward the development of these programs (Dieorio and Miller,2016). Recent publications have sought to clarify the distinction of coaching from advising and mentoring(Dieorio *et al*., 2021).

Coaching for the purposes of this study is defined as a way of learning and aims at transformation, driven by the person’s choices. With its base in a professional relationship of mutual trust and respect, coaching focuses on the person’s aspirations and goals; and works to facilitate their achievement. Coaching embraces provocative and appreciative inquiry, which leads to new insights and shifts. The International Coaching Federation (ICF) defines coaching as partnering with clients in a thought-provoking and creative process that inspires them to maximize their personal and professional potential(ICF, www.coachingfederation.org).

The effects of coaching during medical school are beginning to be studied, measured and appreciated (Wolff *et al*.,2020)(Lovell 2018)(Jasinsk *et al*.,2020). There is support in the literature for the well-being of physicians from coaching (Dyrbye *et al*,2019). A pilot study, performed by Fried and Irwin with coaching for undergraduate students in Canada, measured perceived stress, anxiety and depression, and found significant changes in stress reduction and qualitative support for the positive experience of coaching (Fried and Irwin, 2016).

For the purpose of assessing the benefits of coaching with medical students, a pilot study providing coaching for students throughout their third year of medical school was performed. The aim of this yearlong coaching pilot was to assess how coaching might affect the mental health (stress, anxiety and depression), self-awareness and overall wellbeing for medical students in their third year, during which they are making critical decisions and anticipating their futures. The results might inform the future use of coaching as an integrated part of the educational experience to support students in a meaningful way as we prepare them to embark on their professional training beyond medical school.

## Methods

The Duke University Health System Institutional Review Board for Clinical Investigations conducted a review of this study and declared it to be Exempt from further review in September, 2019 **(Protocol ID:** Pro00103592). Twelve third-year medical students (10% of the cohort) were recruited from two emails sent in September and January to the third-year class explaining coaching, the pilot project and offering eight coaching sessions to each interested student. The email described the definition of coaching, its purpose, procedures and the assessment measures used in this study. The 30-60 minute sessions were spaced at least monthly, with modifications made for individual student schedules. As per coaching protocol, the students selected the topics for coaching discussion at each session and determined the goals for that session. Sessions were initially held in person (ten students) or via Zoom (two students out of the country), but ultimately all transitioned to Zoom due to the COVID pandemic of 2019/2020. The same coach provided all coaching sessions. This coach completed the academic training of the International Coaching Federation (ICF) and earned the credential of Associate Certified Coach. The ICF approved Competencies, Model and the Code of Ethics for ICF framed the practice of this coach. Figure 1 provides the model of coaching that guided the coaching sessions.

**FIGURE 1.**
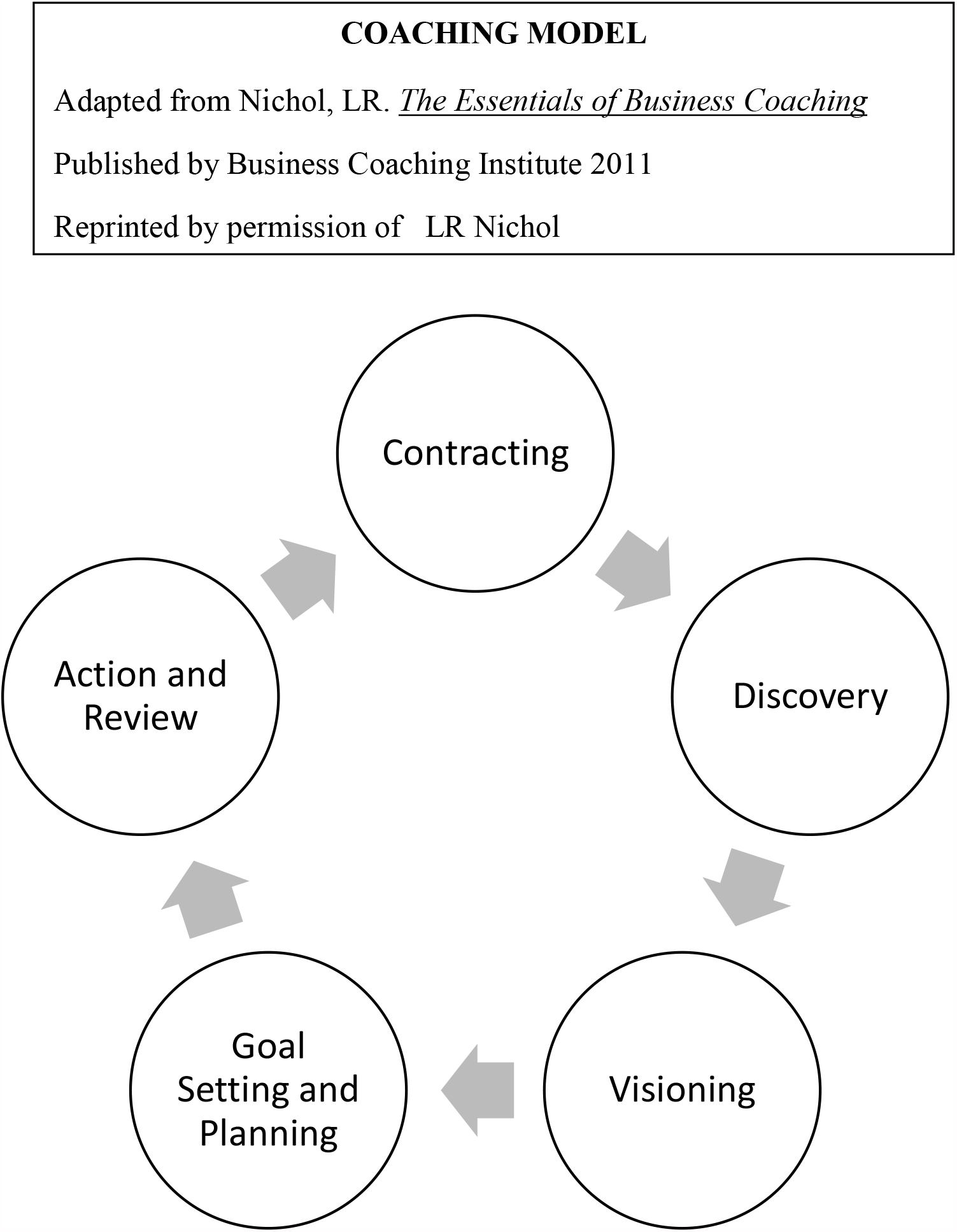

All responding participants signed a consent form and no demographic data was collected outside of gender. A Readiness Assessment for Coaching, adapted by the researchers, with permission from the developer (Gordon-Jones, S), was used to determine the students’ receptivity to coaching. This assessment has 10 items to which the medical student selected yes or no, to determine readiness to participate in coaching. Scores of 9-10 indicate Definitely Ready for coaching; 7-8 indicate Ready for Coaching; 4-6 indicate Coachable with areas to clarify; and 1-3 indicate Not Ready for Coaching. Students needed scores of seven or higher to be eligible for participation in the pilot. There is no reliability and validity data for this assessment.

Each participant, using a self-generated unique identification code to insure de-identification of the data, took the Hospital Anxiety and Depression Scale (HADS) and the Perceived Stress Scale (PSS) pre-, mid-and post-coaching. These scales are easy to administer and are reliable, validated tools(Bjellanda *et al*.,2002)(Aseri and Zohair, 2015)(Measurement Instrument Database for the Social Sciences, www.midss.org)(Cohen *et al*.,1983)(Zigmond and Snaith, 1983). The HADS was devised by Zigmond and Snaith to measure anxiety and depression in a general medical population of patients and includes fourteen questions (seven to assess anxiety and seven to assess depression) that are scored from 0-3 indicating the extent to which a student experiences the feeling in the question (Zigmond and Snaith, 1983). Separate scores for anxiety and depression are calculated, with higher scores indicating more anxiety or depression. The Perceived Stress Scale (PSS) was developed by Cohen et al to measure the degree to which situations in life are appraised as stressful and consists of fourteen questions on a 0(Never)-4(Very Often) point scale, with higher scores indicating more stress (Cohen *et al*.,1983).

Additionally, at the conclusion of coaching, there were three open-ended questions about the coaching experience and a scale to indicate the likelihood of accessing coaching in the future, as shown in Table I.

**TABLE 1.**
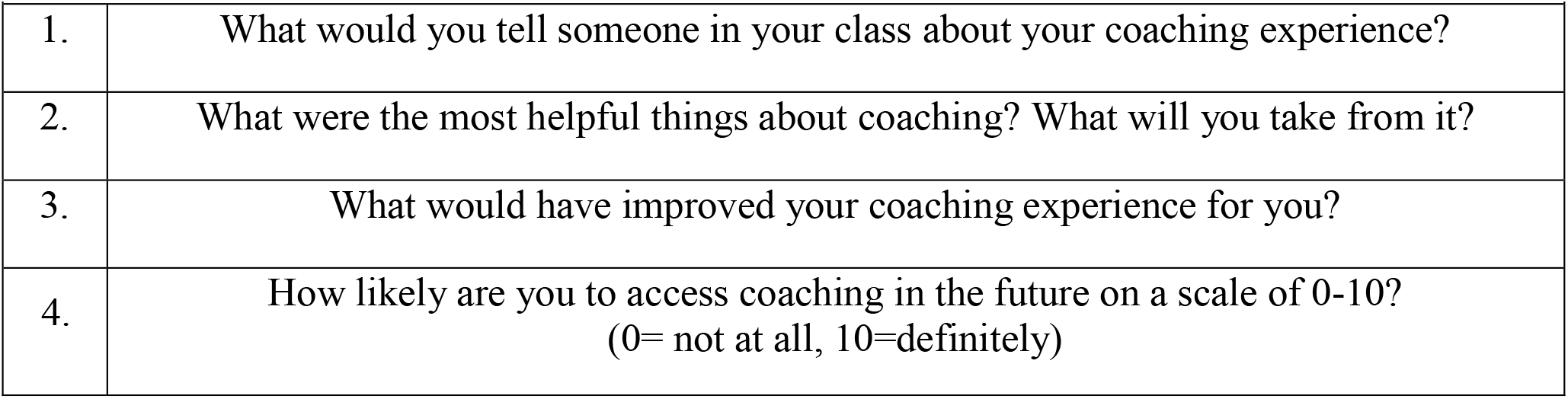
POST-COACHING QUESTIONS.

The HADS and the PSS were administered in hard copy and the open-ended questions were collected via a Qualtrics online survey. Data were analyzed in aggregate and individually for each student utilizing a one-way ANOVA with repeated measures and a Tukey’s post hoc test. The qualitative data were analyzed for emerging themes by consensus of two authors using an applied thematic analysis approach (Guest *et al*.,2012). All qualitative responses were reviewed by each of the two authors independently and discussed together to determine consensus of the underlying focus of each phrase. Themes emerged and were clarified through consensus.

## Results

Of the twelve medical students, eleven identified as female and one identified as male.

Two students were out of the country, so coaching was virtual throughout their experience, whereas ten students transitioned to meeting virtually midway through the coaching. We had 100% response rate for all assessments.

There was a significant effect of coaching on perceived stress. Average PSS scores decreased from 16.92 to 12.25 from pre-test to mid-test, F (1.497, 16.47) = 5.340, p=.023). After four coaching sessions (Mid), PSS scores were significantly lower than baseline (p=.006), as indicated in Figure 2.

**FIGURE 2.**
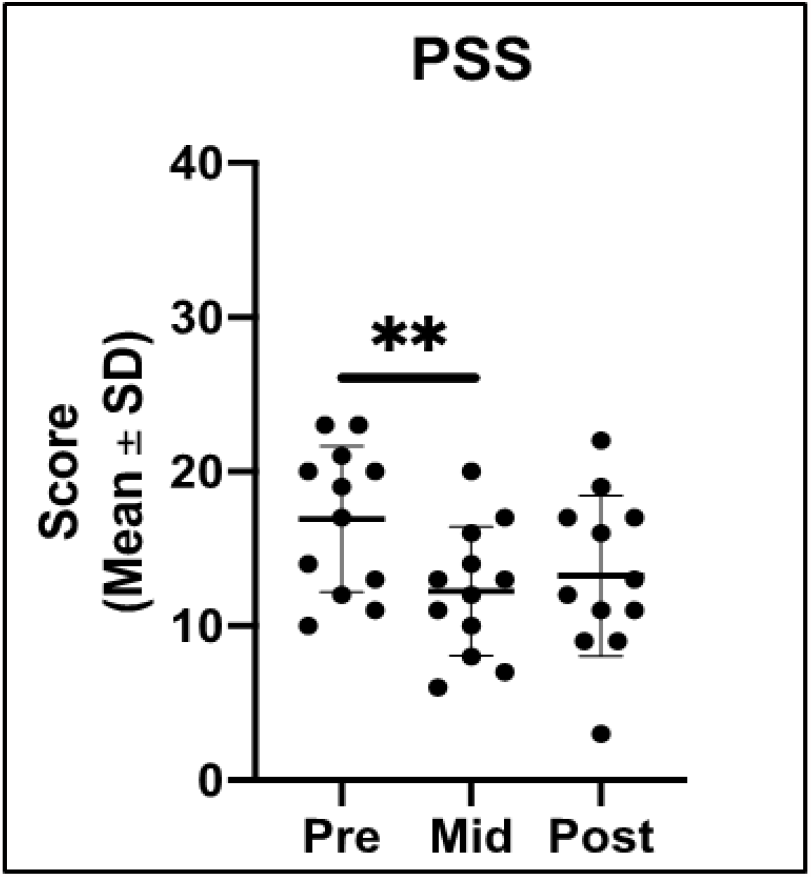
Perceived Stress Scale Measures.

There was a trend toward a significant effect of coaching on anxiety from pre- to mid-test (HADS-Anxiety; F (1.957, 21.53) = 3.407, p=.053), as indicated in Figure 3. However, from pretest to mid-point in the coaching the HADS-A scores decreased from 8.25 to 6.17, which did not reach significance (p=.079).

**FIGURE 3.**
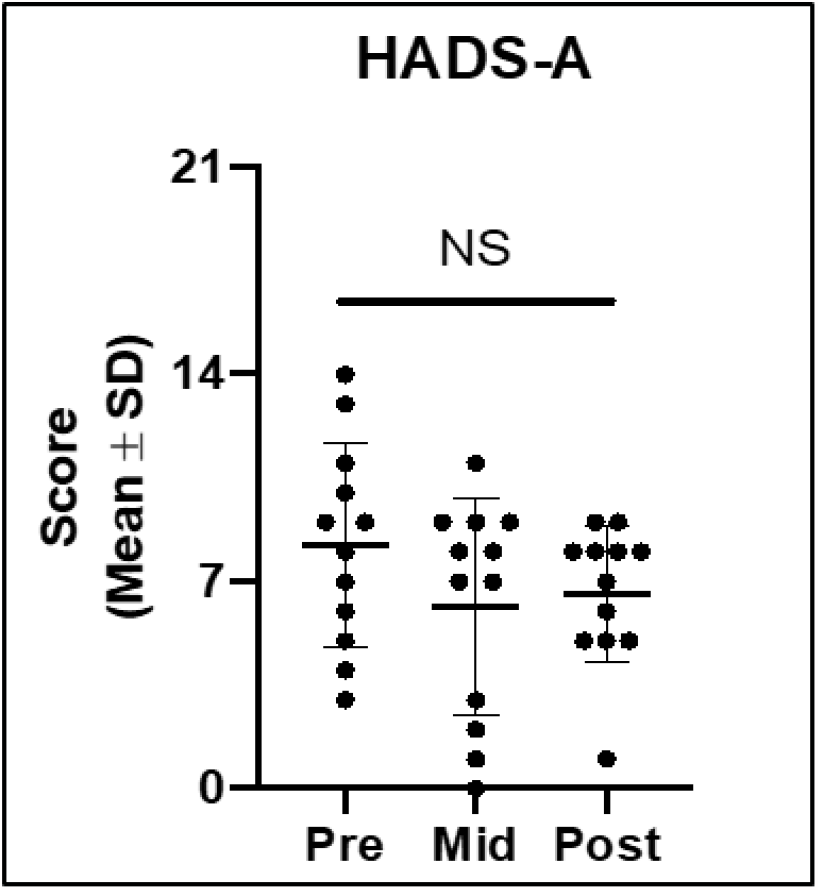
Hospital Anxiety and Depression Scale Anxiety Measures.

There was no significant effect of coaching on depression (HADS-D; p>.05), as shown in Figure 4.

**FIGURE 4.**
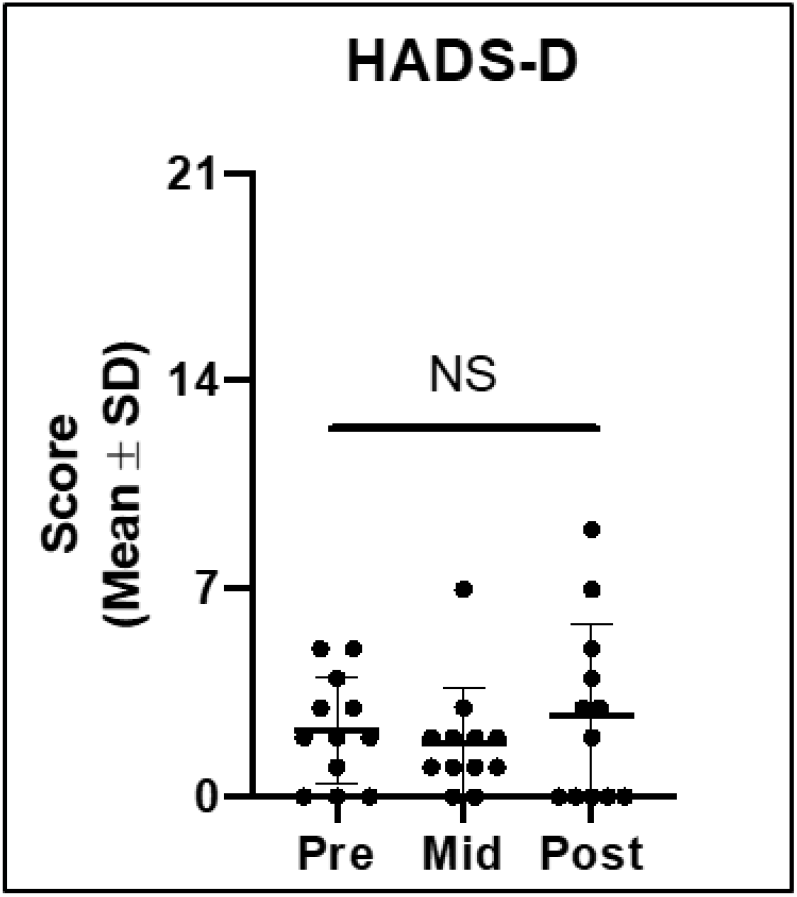
Hospital Anxiety and Depression Scale Depression Measures.

The measured changes from pre- to post coaching and mid- to post-coaching in PSS and HADS-A and HADS-D were not significant.

The qualitative analysis revealed responses in four domains: 1) Affective, 2) Cognitive, 3) Skills and 4) Attributes of the Coaching Experience. Affective responses included gaining perspective, self-awareness and positive feelings on the part of the student. Cognitive responses included goal setting and working through solving problems. Students indicated that the coaching helped them set both personal and professional goals. The Skills responses included developing reflection abilities and critical thinking.

Attributes of the coaching experience included perceiving coaching as a positive, individualized and supportive experience. In the open-ended questions after coaching, students identified strengths and areas for improvement of their coaching experience. Students found their coaching experience would have been improved by meeting more frequently, meeting in person, meeting in a more private location and starting coaching earlier in medical school.

A wide variety of topics were chosen by the students for coaching, including choice of field for residency, residency program attributes and applications, assertiveness skills, delegation skills, work-life balance, self-care and boundaries in healthcare and life, among others. Many expressed that the questions asked by the coach had not been considered previously and facilitated self-reflection in planning for the future.

Examples of these responses are provided below in Table 2.

**TABLE 2.** QUALITATIVE RESPONSES EXAMPLES

|  |  |  |
| --- | --- | --- |
| **AFFECTIVE RESPONSES** | **“This experience made me more confident in myself and equipped me with the tools to address and conquer any stressor or source of anxiety facing me in my professional life”** | **“To be able to share freely and be asked open-ended questions where I could problem-solve and think through my challenges was empowering.”** |
| **COGNITIVE RESPONSES** | **“The coaching experience was able to reflect back to me my thoughts, feelings and concerns in a way that was just different enough to give me insight and a new perspective...motivated me to pursue my own definition of success... will push me to ask myself what the next steps would be to achieve what it is I am working on”** | **“Perhaps the most useful thing for me was goal setting and then discussing what steps need to be taken to get there.”** |
| **SKILLS RESPONSES** | **“I was able to walk away from each session with actionable items and the sense that I was working on ways to handle whatever issues I had discussed in the meeting...helped me feel more in control of my situation or at least with tools to handle a situation in ways I wanted to.”** | **“My coach helped me think about how I can most effectively explore my interests. She helped me think critically about how I approach problems in my life. These are skills I will take with me.”** |
| **ATTRIBUTES OF COACHING EXPERIENCE** | **“I loved having someone from a different perspective to talk to me about strengths and challenges. It was just an invaluable resource”** | **“Everyone can benefit from a coach and coaching is flexible and customizable”** |
| **STRENGTHS OF COACHING** | **“I enjoyed the safe space created for more abstract discussions on purpose, meaning and motivation. It helped make the experience about self-transformation as much as it did about setting and achieving goals”** | **“Coaching is a wonderful tool that helps me work out any issues or focus on any personal development I want to do. It is more personal than advising, but more professional than therapy.”** |
| IMPROVEMENTS FOR COACHING EXPERIENCE | **“Having coaching opportunities from the beginning of medical school would have been extremely helpful in my development over these formidable years. While 8 sessions were better than no sessions, having an even longer longitudinal experience would have been great.”** | **“Although we had a limited number of sessions, it would have been great to meet more frequently as it allows for greater accountability with goals and closer check-ins.”** |
| VARIETY OF TOPICS DISCUSSED | **“I appreciated that we covered every topic that I wished to. Having never been coached before (outside of sports), I was fascinated by the approach”** | **“Whether it was with my research, my plans for residency, or lifestyle goals we always discussed how to break it down into manageable elements, manage time, and deal with challenges that arise.”** |

When asked about their likelihood of seeking future coaching, 92% (11/12) of respondents reported being likely to seek coaching in the future (score > 5) with 75% (9/12) indicating high likelihood (score > 7). The mean likelihood score was 8.3/10 with a median score of 8.5/10 and a range of 5.

## Discussion

Our data reveal that coaching appears to have positive effects on stress reduction and anxiety reduction after four of the eight sessions (mid-point in the experience). Our data match the findings found in the Fried and Irwin study in that the mid- to post-results were not significant. (Fried and Irwin, 2016). The onset of COVID 19 restrictions that occurred midway through the coaching sessions required a change to virtual format for all students, which may have influenced the effects measured at the end of the sessions. The introduction of major changes and stresses in the lives of these students, resulting from the pandemic, may have had effects on our data and results. The lack of significant change from mid- to post coaching in our measures might indicate the effects of COVID 19 restrictions or may indicate that four sessions are optimal for achieving the changes we measured. The depression measures were not affected by the coaching, which is not surprising given that coaching is not intended as an intervention for depression.

Student responses revealed meaningful effects on their affective, cognitive and skill domains, as well as positive personal experiences from coaching. The qualitative themes we found echo those of the Fried and Irwin study, which were managing stress, self-awareness, appreciating different perspectives and self-reliance. (Fried and Irwin, 2016). A recent study of coaching with practicing physicians revealed that coaching reduced emotional exhaustion and symptoms of burnout, thus improving the quality of life and resilience for those physicians. (Dyrbye *et al*,2019) The range of topics chosen by students for coaching highlights the broadness of coaching as resource for these students. Coaching has potential for providing medical students with a positive experience and support to navigate the stresses and anxiety inherent in the medical school journey and beyond.

The limitations of this study that compromise its generalizability are the small sample size, the lack of a control group, limited demographic data collected, the lack of reliability and validity data for the Readiness Assessment tool and the students coming from one specific program and institution. The provision of coaching by a single coach in this study may be seen as a strength, such as consistency of skill, or limitation, due to being an unrealistic expectation in large-scale implementation of coaching. Future study with more students in different programs at different institutions, inclusion of a control group and more demographic data will add more evidence about the effects of coaching with medical students.

## Conclusions

Coaching holds promise as an intervention that could be offered to medical students to reduce stress and anxiety, as well as provide support for affective, cognitive and skills development, preparing them for their professional futures.

## Data Availability

Data is available from the corresponding author. There are no external datasets

## Declarations

### Ethics approval and consent to participate

The Duke University Health System Institutional Review Board for Clinical Investigations has conducted a review of this study and declared it to be Exempt from further review in September, 2019 **(Protocol ID:** Pro00103592).

### Authors Contributions

ER designed the study, provided the intervention, drafted and revised the manuscript, provided final approval and agreed to be accountable for all aspects of the work. TF contributed the analysis and interpretation of the data, revised, provided final approval of the manuscript and agreed to be accountable for all aspects of the work. KC assisted with the design of the study, assisted with data collection, analysis and interpretation, revised, provided final approval of the manuscript and agreed to be accountable for all aspects of the work.

## Acknowledgements

The authors express gratitude to Sackeena Gordon-Jones for allowing them to adapt and utilize the Readiness Assessment for Coaching in this study. Additionally, the authors wish to express appreciation to Marilyn H. Oermann, PhD, RN, ANEF, FAAN, Duke School of Nursing, for her review of this manuscript and Duke AHEAD for funding and the Grant Committee for encouraging this project.

## REFERENCES

1. Aseri A, Zohair A. Reliability and validity of the Hospital Anxiety and Depression Scale in an emergency department in Saudi Arabia: a cross-sectional observational study. BMC Emerg Med. 2015;15(28). doi: 10.1186/s12873-015-0051-4

2. Bjellanda I, Dahlb AA, Haugc TT, Neckelmannd D. The validity of the Hospital Anxiety and Depression Scale: An updated literature review. Journal of Psychosomatic Research 2002;52: 69–77

3. Cary M, Penner J, Mikhaiel JP. (STAT)Coaching and leadership training can help med students avoid burnout. March 25, 2019. https://www.statnews.com/2019/03/25/coaching-leadership-training-avoid-burnout-medical-students/

4. Cohen S, Kamarck T, Mermelstein, R. A global measure of perceived stress. Journal of Health and Social Behavior. 1983;24(4), 385–396. doi: 10.2307/2136404

5. Deiorio, N, Miller Juve, A. Developing an academic coaching program. doi: https://doi.org/10.15694/mep.2016.000143 Published Date: 07/12/2016

6. Deiorio NM, Hammoud MM. Coaching in Medical Education: A Faculty Handbook. 2017. https://www.ama-assn.org/education/accelerating-change-medical-education/coaching-medical-education-faculty-handbook

7. Deiorio Nicole M. MD; Foster Kenneth W. EdD; Santen Sally A. MD, PhD Title: Coaching a Learner in Medical Education DOI: 10.1097/ACM.0000000000004168

8. Dyrbye LN, Thomas MR, Shanafelt TD. Systematic Review of Depression, Anxiety, and Other Indicators of Psychological Distress Among U.S. and Canadian Medical Students. Academic Medicine. 2006;81(4): 354–373.

9. Dyrbye, LN, Shanafelt, TD, Gill PR, Satele BA, West CP. Effect of a Professional Coaching Intervention on the Well-being and Distress of Physicians. A Pilot Randomized Clinical Trial. JAMA Intern Med. 2019. doi:10.1001/jamainternmed.2019.2425. Published August 5, 2019.

10. Fried, RR, Irwin, JD: Calmly coping: A Motivational Interviewing Via Co-Active Life Coaching(MI-VIA-CALC) Pilot Intervention for University Students with Perceived Levels of High Stress. International Journal of Evidenced Based Coaching and Mentoring. 2016;14(1):16–33.

11. Guest G, MacQueen K, Namey E. Applied Thematic Analysis[Internet]. 24455 Teller Road, Thousand Oaks California 91320 United States: SAGE Publications, Inc.; [accessed 2021 Jan 25]. http://methods.sagepub.com/book/applied-thematic-analysis

12. (ICF) Ethics, Model, Competencies. Accessed May 1, 2021 www.coachingfederation.org

13. Jasinski, J.C., Jasinski, J.D., Härtel, C.E.J. and Härtel, G.F. (2020), “Development and Evaluation of an Online Coaching Model for Medical Students’ and Doctors’ Mental and Physical Well-being Management”, Härtel, C.E.J., Zerbe, W.J. and Ashkanasy, N.M. (Ed.) Emotions and Service in the Digital Age (Research on Emotion in Organizations, Vol. 16), Emerald Publishing Limited, Bingley, pp. 69–93. https://doi.org/10.1108/S1746-979120200000016007

14. Kumar B, Shah MAA, Kumari R, Kumar A, Kumar J, Tahir A. Depression, Anxiety, and Stress Among Final-year Medical Students. Cureus. 2019 ;11(3):e4257.

15. Lovell, B. What do we know about coaching in medical education? A literature review. Med Educ. 2018;52(4):376–390. doi: 10.1111/medu.13482. Epub 2017 Dec 11.

16. Measurement Instrument Database for the Social Sciences. Retrieved from March, 2021 https://www.midss.org/content/perceived-stress-scale-pss

17. Wolff, M, Hammoud M, Santen S, Deiorio N, Fix M. Coaching in undergraduate medical education: a national survey. Med Educ Online. 2020 Dec;25(1):1699765. doi: 10.1080/10872981.2019.1699765

18. Zigmond AS, Snaith RP. The Hospital Anxiety and Depression Scale. Acta Psychiatr Scand 1983;67: 361–370.

